# Warmer weather and global trends in the coronavirus COVID-19

**DOI:** 10.1101/2020.04.28.20084004

**Authors:** Hong Li, Hongwei Xiao, Renguo Zhu, Chengxing Sun, Cheng Liu, Hua-Yun Xiao

## Abstract

Predicting COVID-19 epidemic development in the upcoming warm season has attracted much attention in the hope of providing helps to fight the epidemic. It requires weather (environmental) factors to be included in prediction models, but there are few models to achieve it successfully. In this study, we proposed a new concept of environmental infection rate (*R*_E_), based on floating time of respiratory droplets in the air and inactivation rate of virus to solve the problem. More than half of the particles in the droplets can float in the atmosphere for 1–2 hours. The prediction results showed that high *R*_E_ values (>3.5) are scattered around 30°N in winter (Dec.-Feb.). As the weather warms, its distribution area expands and extends to higher latitudes of northern hemisphere, reaching its maximum in April, and then shrinking northward. These indicated that the spread of COVID-19 in most parts of the northern hemisphere is expected to decline after Apr., but the risks in high latitudes will remain high in May. In the south of southern hemisphere, the *R*_E_ values tend to subside from Apr. to July. The high modeled *R*_E_ values up to July, however, suggested that warmer weather will not stop COVID-19 from spreading. Public health intervention is needed to overcome the outbreak.

## 1. Introduction

The COVID-19 pandemic, caused by severe acute respiratory syndrome coronavirus 2 (SARS-CoV-2), has sickened >2.2 million people and killed >0.15 million people in 202 countries worldwide as of 18 April 2020, and may greatly weaken the global economy^5^. Numerous studies on past outbreaks such as influenza^6–8^ showed that spread of respiratory infectious diseases seems to be easier at low temperatures and low humidity. So more and more people and governments hope that warmer weather will slow down or stop the spread of COVID-19. But there is still no evidence to support it^9^.

Unlike diseases that rely on insect vectors for transmission, for example malaria^10^, predicting human-to-human respiratory diseases such as COVID-19 is somewhat difficult. A key parameter for model prediction is the basic reproduction rate (*R*_0_), which is usually estimated with various types of complex mathematical models. The *R*_0_ value is affected by biological (cell structure^11^ and concentrations of virus, immunity of susceptible population, etc), socio-behavioral^12,13^ and environmental factors^14^. Because environmental factors are different in regions and time, it may be unsuitable for the applicability of *R*_0_ outside the region where it was calculated^15^. Actually, many studies have found close correlation of epidemic development with environmental factors such as wind, temperature *(T)* and relative humidity *(RH)* ^6–8^, which affects viable virus concentrations and exposure time of virus to susceptible population. Here, a new reproduction rate that depends on environmental factors (*T* and *RH)*, environmental reproduction rate (*R*_E_), was introduced. Using *R*_E_, it is possible for us to predict the epidemic development with time (season) and places (countries) of a specific respiratory disease.

## 2. Methods

### 2.1 Estimation of *R*_E._

In the *R*_E_ estimation, we assumed that there was no individual difference (age and gender) in the COVID-19 transmission^16^ because biological structure of SARS-CoV-2 is suitable for infecting all people^11^ and SARS-CoV-2 is a new virus, almost all people have no immunity to the disease^1^. The *R*_E_ value of COVID-19 can be estimated with effective floating time of respiratory droplets in the air (*t*_C0_) (Fig. 1), the infection period *(d*_i_) and a correlation coefficient *(β)* in the absence of control measures. The *t*_C0_ is a factor that only correlated with meteorological parameters (*T* and *RH*) and half lifetime (*t*_1/2_) of active viruses in aerosols.

**Figure 1.**
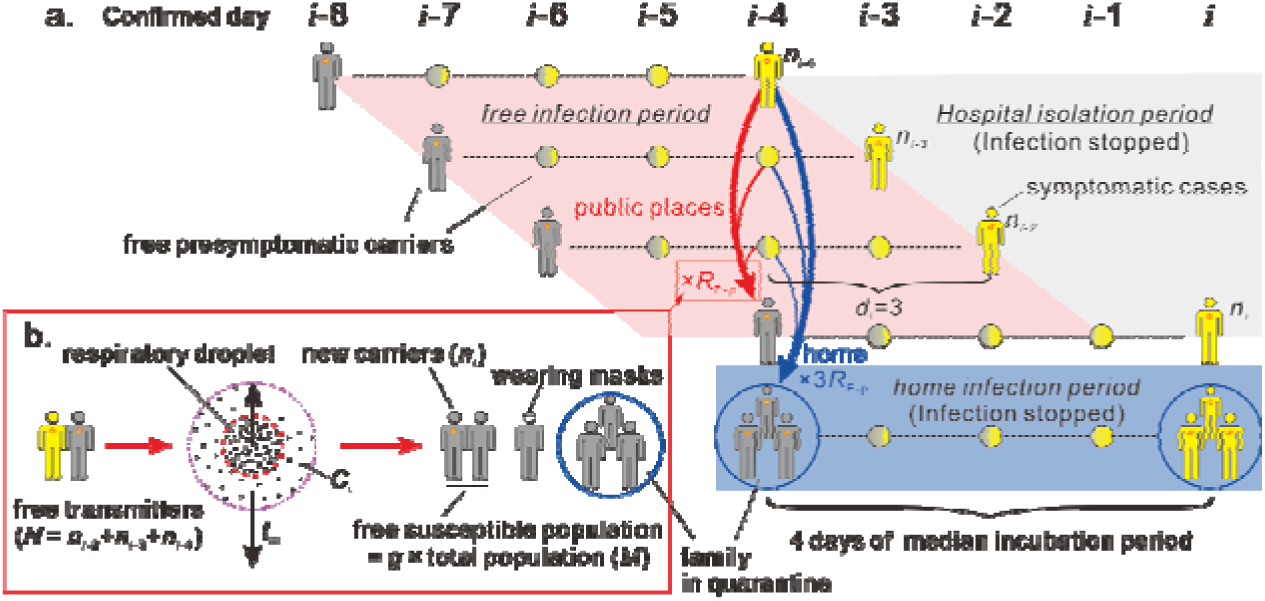
Infection mode of COVID-19. a. Two ways of infection. New cases (*n*_i-4_) and carriers 48 hours before onset *(n*_i_*_-_*_2_ and *n*_i-3_) are contagious to family members (blue arrows) and susceptible people (*n*_i_) in public places (red arrows). The incubation period and infection period (*d*_i_) were set at 4 days^23^ and 3 days, respectively. b. Conditions for spreading viruses in public places. See Method Section for more details.

The concentration of viable viruses in droplets at the time *t* depends on the inactivation rate (*α*), which can be described by^17^

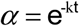

where *k* is the rate coefficient of inactivation. When *α* is 0.5, *t* is the half lifetime of the virus (*t*_1/2_, min). The half lifetime of SARS-CoV-2 is 1.1 hours^18^, and then *k* can be calculated to be 0.0105. So

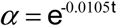

The volume of the droplet air mass changes with air temperature, which will affect the concentrations of viruses in the air. Using the Ideal Gas Law, the concentration of virus in droplets *(C_T_)* at air temperature *T*(°C) is

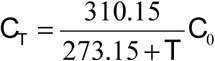

where *C*_0_ and 300.15K are the initial concentration of viruses and temperature in the mouth or nasal cavity. So the concentration *(C_t_*) of viable viruses at the time *t* is

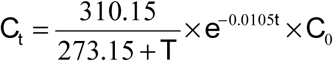

Then the effective floating time *(t*_C0_, hours) of virus-bearing droplets with *C*_0_ is

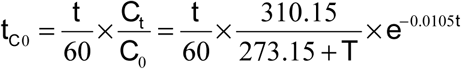

In the presence of control measures,

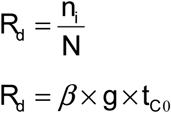

where *g* is the proportion of susceptible population; *n*_i_ is the number of new COVID-19 cases confirmed at day *i*; *N* is the sum of free COVID-19 transmitters confirmed at days *i*-4, i-3 and *i*-2 (Fig. 1a, 1b); *β* (h^-1^) is the correlation coefficient between *g×t*_C0_ and *R*_d_. In the absence of control measures *(g* = 1), environmental reproduction rate (*R*_E_) can be calculated by

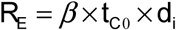

where *d*_i_ is the infection period of COVID-19. In the study, *d*_i_ is 3 days. So

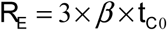

When *RH* is less than 50%, the equilibrium particle size (*d*_eq_) of droplets will not change with *RH* (Fig. 2). Therefore, in the mathematical model established, we regarded *RH* less than 50% as 50%.

**Figure 2.**
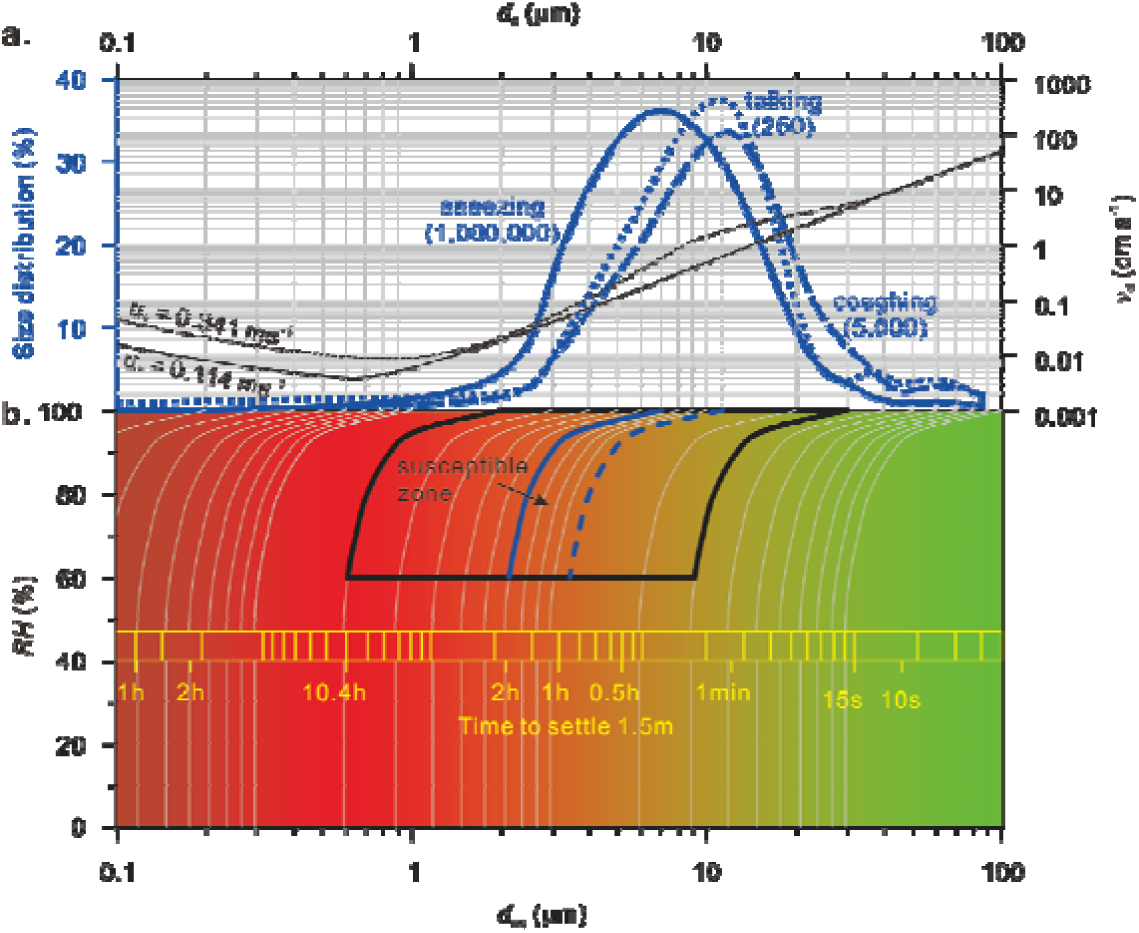
Control of relative humidity on the floating time of droplets in the air. a. Size distribution and deposition velocities (*v*_d_). The figure in brackets is the number of droplet particles. Size distribution and *v*_d_ are cited from refs 18 and 24. The u* is the friction velocity. b. Equilibrium particle size (*d*_eq_) and floating time of droplets under different *RH* conditions. The black and blue line ranges represent the susceptible areas caused by 90% and peak number of droplets, respectively.

### 2.2 Data availability

The number of confirmed cases of coronavirus disease 2019 (COVID-19) in all countries, except China, is from Johns Hopkins CSSE (https://www.arcgis.com/apps/opsdashboard/index.html#/bda7594740fd40299423467b48e9ecf6,). The number in Chinese provinces except Hubei is from Chinese Center for Disease Control and Prevention (http://2019ncov.chinacdc.cn/2019-nCoV/,). The number in Hubei is from National Health Commission of the People’s Republic of China (http://www.nhc.gov.cn/xcs/yqtb/list_gzbd.shtml). The number of the onset in Wuhan is from Report of the WHO-China Joint Mission on Coronavirus Disease 2019 (COVID-19).

The data of monthly and 10-days average global surface temperature (*T*, °C) and relative humidity *(RH*, %) is from Geospatial Interactive Online Visualization and Analysis Infrastructure (Giovanni, https://giovanni.gsfc.nasa.gov/giovanni/, AIRS/Aqua L3 Daily Standard Physical Retrieval (AIRS-only) V006 1 °C × 1 °C). The ground station data of temperature and relative humidity is from ground observation data (http://www.weatherandclimate.info and https://www.wunderground.com/history).

Population data leaving Wuhan is from https://qianxi.baidu.com/. Population grouped by age in China, Guangzhou and Wuhan are from Chinese National Bureau of Statistics (http://www.stats.gov.cn/tisj/ndsj/. 2017), Guangzhou Statistics Bureau (http://tjj.gz.gov.cn/pchb/dlcrkpc/, the sixth census) and Wuhan Statistics Bureau http://tjj.wuhan.gov.cn/newslist.aspx?id=2012111010461248, 2017). respectively.

## 3. Results

The *RH* controls the floating time of respiratory droplets in the air. A longer floating time of virus-bearing droplets in the air will increase the risk of exposure to the virus. Droplets do not evaporate completely in the air because they contain substances such as salts and proteins^19^. The number of droplet particles at 2–12 microns (μm) ejected during speaking. coughing. sneezing accounts for more than 90%. and the initial particle size *(d*_0_) of the peak number is 7–10μm^20^ (Fig. 2a). Droplet particles of large size (>100μm) settled rapidly due to gravity. The droplet particles of small size reached an equilibrium particle size (*d*_eq_) rapidly (in a few seconds^14,19^) and floated in the air for some time (t) depending on *RH* (Fig. 2b). When *RH* is <95%. the main natural weather conditions. the settling of >90% droplet particles may take more than 5 hours (Fig. 3). For 10–50μm (*d*_0_) droplets. the exposure risk under low *RH* (60%) is more than 20 times larger than that at *RH* 100% (Fig. 4). The 7–10μm (*d*_0_) droplets. floated in the air for up to 1–2 hours under *RH* 60%. are the most important vector of virus transmission (Fig. 5a). Here we used the floating time of 10μm droplets in the *R*_E_ estimation. When *RH* is in the range of 50–100%. the floating time (*t*, min) of droplets with initial particle size (*d*_0_) of 10μm in the air is a function of *RH* (Fig. 5a)

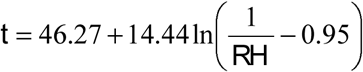

**Figure 3.**
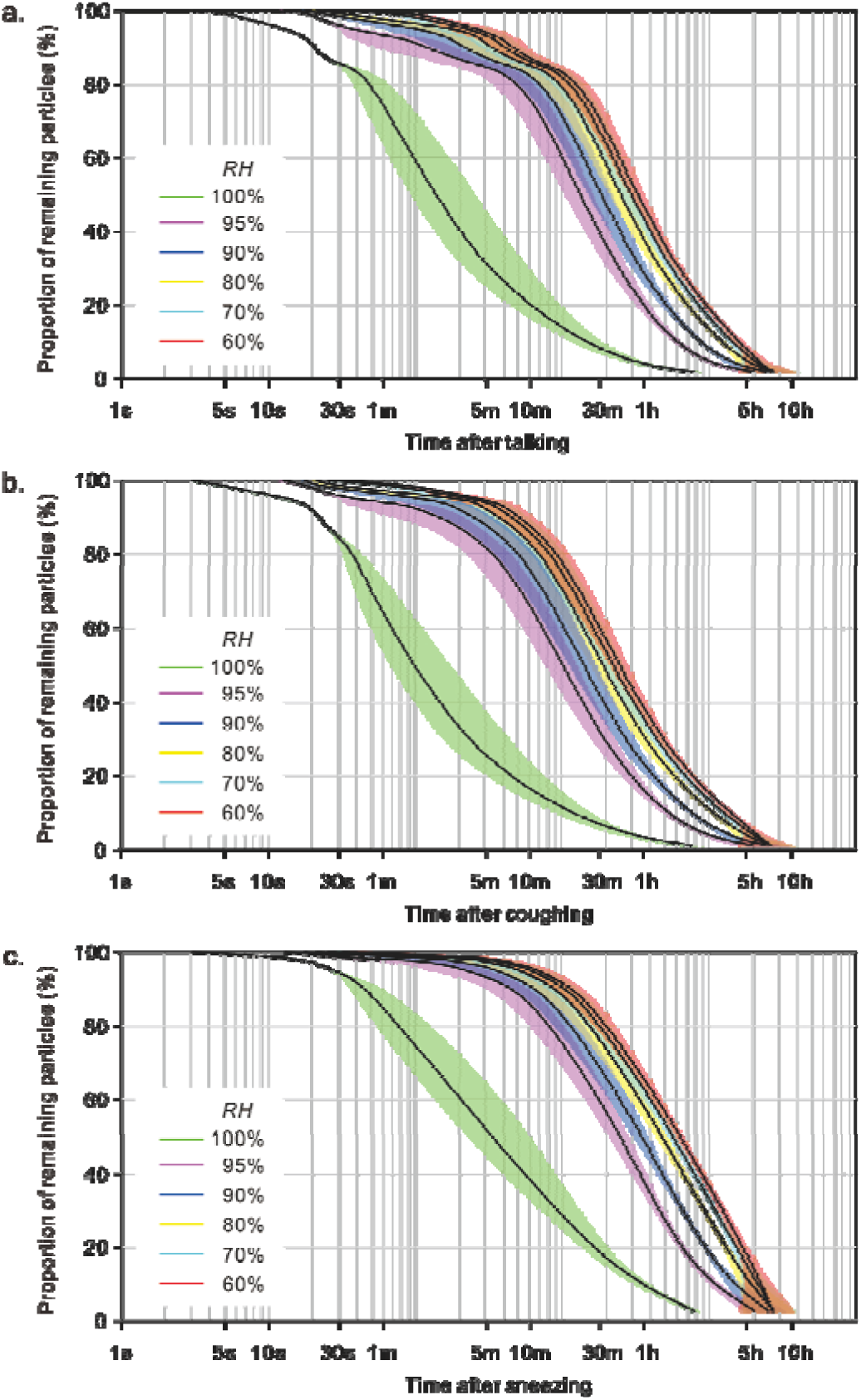
Effect of *RH* on the relationship between the floating time of droplets in the atmosphere and the proportion of remaining particles. a. Speaking. b. Coughing. c. Sneezing.

**Figure 4.**
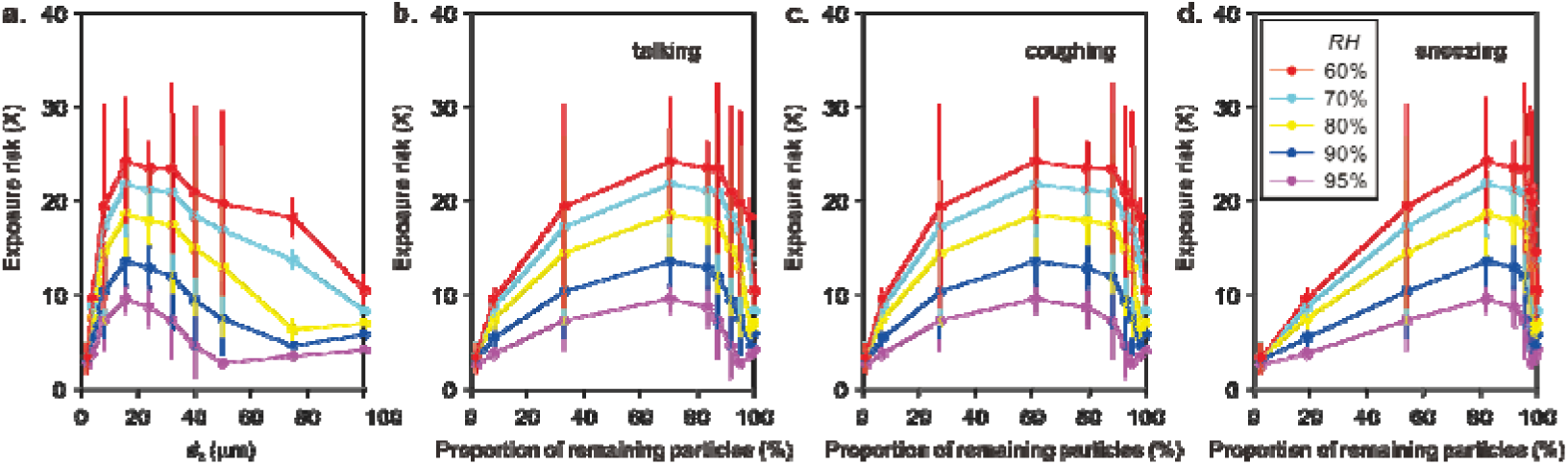
Increasing exposure risk to droplets due to lower *RH*. a. d0. b. Speaking. c. Coughing. d. Sneezing. The risk of exposure is expressed by dividing the float time of RH100%.

**Figure 5.**
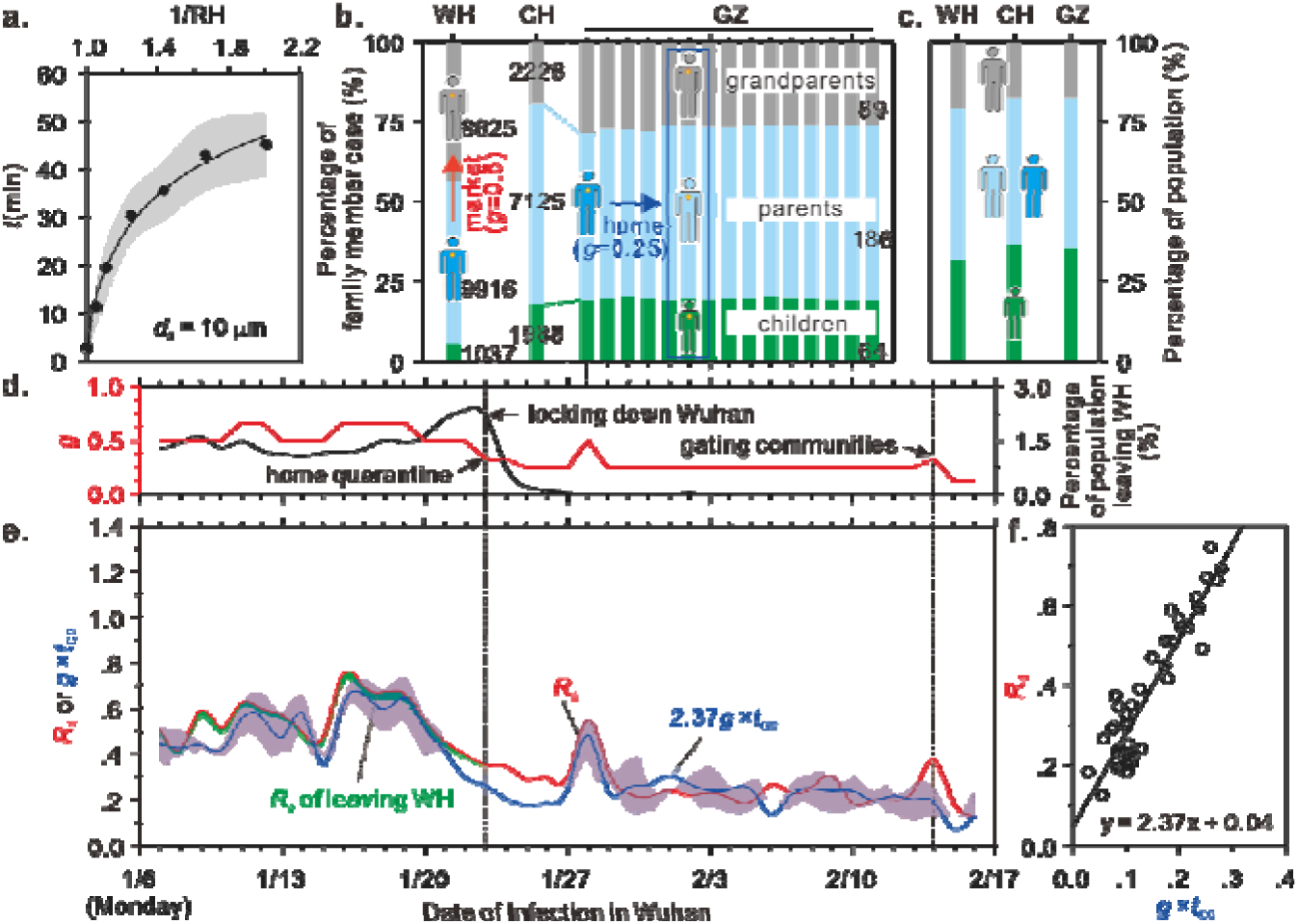
Modeling the COVID-19 epidemic development in Wuhan. a. The relationship between floating time (t) of 10μm droplets in the air and *RH (r* = 0.99, *p* < 0.001). Data are calculated from Fig. 2. Shaded region is calculated with u* of 0.114 and 0.341 m s^-1^. b. Age structure and family member structure of confirmed cases in China, Wuhan (WH) and Guangzhou (GZ) during the outbreak. The figure in brackets is the number of cases. The date of infection is equal to the date of confirmation minus 14 days for WH^21^ and 5 days for GZ. c. Age structure of population in China (2017), WH (2017) and GZ (2011). d. Effect of control measures on the proportion of susceptible population (g) and the proportion of people leaving WH. e. Daily infection rate in public places (*R*_d_) and modeled *g× t*_C0_ values. The red line and its shaded region are calculated with a 4-days incubation period and 2 to 7-days incubation periods^23^. Shaded region of *g×t*_C0_ marks the 95% prediction envelops. f. Relationship between *R*_d_ and modeled *g×t*_C0_ *(r* = 0.92, *p* < 0.001).

People are infected only when the amount of viable viruses exhaled exceeds the minimum infective dose (MID). The inactivation rate (*α*) of viruses in aerosols can change concentrations of viable viruses in the air and then the infection rate. The concentrations of active viruses decreased exponentially in the air, with a *t*_1/2_–related inactivation coefficient (*k*)^19^. The *t*_1/2_ of SARS-CoV-2 in aerosols is 1.1 hours^20^, similar to the floating time of 7–10μm particles in the air under *RH* 60% (Fig. 2a), indicating that ~ half of viruses died during settling from the air in dry weather.

The number of new cases and meteorological parameters during the outbreak in Wuhan (WH), China, have been analyzed, allowing us to fit the *t*_C0_ to daily infection rate in public places (*R*_d_). In the presence of control measures, *β×g×t*_C0_ = *R*_d_, where *g* is the proportion of susceptible population.

The implementation of control measures for public health intervention in WH includes locking down the city (January 23), home quarantine (January 23) and gating communities (February 14). Before January 23, COVID-19 patients were found mainly elderly (>60 years old) and middle-aged (30–60 years old), each accounting for ~50%^21^ (Fig. 5b), and mainly associated with food shopping^22^. So we believed that half of susceptible population (young and half the middle-aged) in the city was not involved *(g* = 0.5). The age structure of cases changed after January 23. For example. according to the epidemiological survey data in Guangdong and Sichuan provinces (China)^16^. among all the 1.836 cases. 1.308 cases were associated with 344 times of aggregation (1 person infected 3 people). and about 3/4 of the cases confirmed were mainly in the family. The age structure of the cases is very consistent with the age structure and family member structure of China and the two provinces (Fig. 5c). Therefore. 3/4 of the susceptible population were isolated at home during this period. i.e. *g* = 0.25. After gating communities on February 14. the number of people who appeared in public places decreased sharply to 1/8 (one person from one family went out on the second day). and thus g was 0.125 (Fig. 5d). Additionally. in the *R*_E_ estimation we increased the *g* value on some days due to special events. e.g. buying food on weekends (custom) before locking down the city. outdoor activities on sunny day of January 28 after a long period of rain and a claim on returning to work on February 14 (canceled on February 13) (Fig. 5d).

The *t*_C0_ (hour) obtained by running the model (See Method Section) is in good agreement (r = 0.92. p < 0.001) with *R*_d_ (Fig. 5e. 5f). meaning that *t*_C0_ corresponds to *R*_d_ of 2.37 every 1 hour. According to this. modeled daily infection rate *(R*_E-d_) during the entire epidemic in WH averaged 0.93(95% CI: 0.50–1.30). Since the *R*_d_ value was calculated with an assumption that *d*_i_ is 3 days. the *R*_E_ value then was 2.8. which is equivalent to the reported *R*_0_ value^3,4^. The 3 days of infection period (*d*_i_) of COVID-19 are considered based on the reports that COVID-19 patients are infectious two days before^16^ and one day after onset of illness (Fig. 1 a). The *d*_i_ value is lower than that reported in ref. 3 (5.2 days). But increasing *d*_i_ means a decrease in the *R*_d_ calculated (Fig. 1b; See Method Section). so the *R*_E_ value does not change much with *d*_i_. After home quarantine on January 23. the *R*_E-d_ averaged 0.25(95% CI: 0.11–0.59) and thus the modeled public infection rate (*R*_E-p_. =*R*_E-d_×*d*_i_) was 0.75. corresponding to a declined COVID-19 spread by 73%. After gating communities on February 14. the mean *R*_E-p_ value dropped to only 0.33. and the disease diminished considerably by 88% (Fig. 5e). indicating that these control measures are very effective to prevent the spread of the epidemic. In the estimation. we did not consider family cases because they were basically isolated at home. and the infection stopped later.

The outbreak of COVID-19 was mainly in China. South Korea. Iran and Italy around 30°N before March 2020. and spread to almost all parts of the world after March 10 (Fig. 6a). In Europe. for example. the epicenter was Italy before March. After the 10th day of March. it raged in Western Europe and began to spread in Northern Europe. Global distribution of *R*_E_ value modeled with 10-day averages of *T* and *RH* showed a similar trend. In the modeled R_E_-distribution maps (Fig. 6b). regions with the *R*_E_ values higher than 3.5 are scattered in Europe in early March. while linked together after March 10. This assured us that meteorological conditions in March played an important role in promoting the rapid spread of COVID-19.

**Figure 6.**
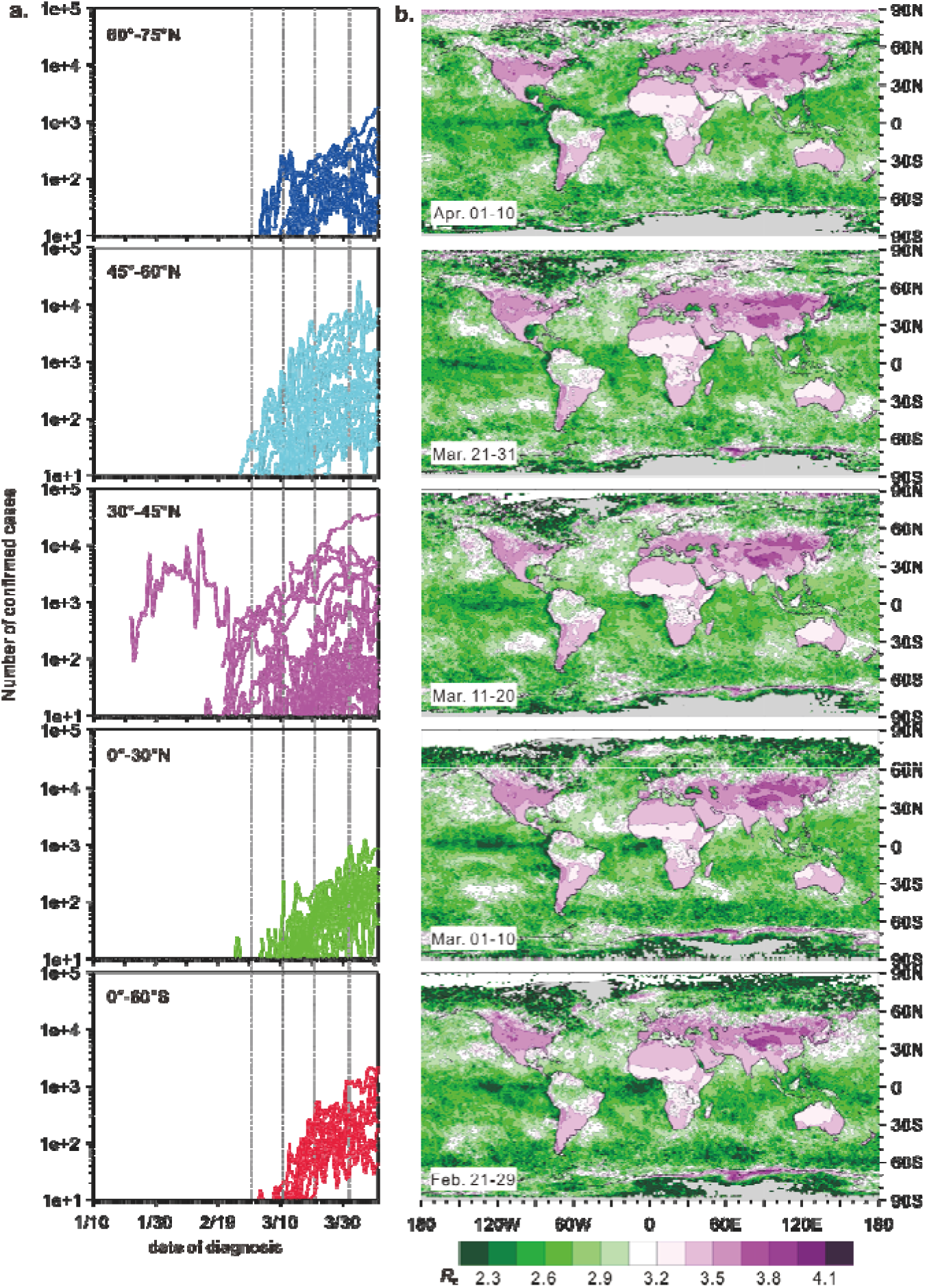
Global COVID-19 epidemic development in different latitudes (see Supplementary Table S1) (a) and 10-days modeled *R*_E_ values (b). Dense lines in panel a can indicate concentrated outbreaks.

The modeled *R*_E_ value of COVID-19 worldwide ranged between 2.15 and 4.25 (Fig. 7). The low *R*_E_ value appears at ocean, and the value on land in temperate zone is generally higher than 2.8 even in July, indicating that warmer weather may not halt the spread of COVID-19 at the stage when humans have no immunity to SARS-CoV-2. In northern hemisphere, distribution areas with high modeled *R*_E_ values (>3.5) are the largest in April and then shrinks to the north. It suggested that in the region of 30°-60°N where the outbreak was very severe in April, warmer weather was conducive to slowing the spread of COVID-19 while the risk in Northern Europe, Russia and Canada is still high in May. In southern hemisphere, regions with high global *R*_E_ values (>3.3) also expand northward as air temperature rises. The fall of *R*_E_ value is the most obvious in East and South Asia and New Zealand in warmer season.

**Figure 7.**
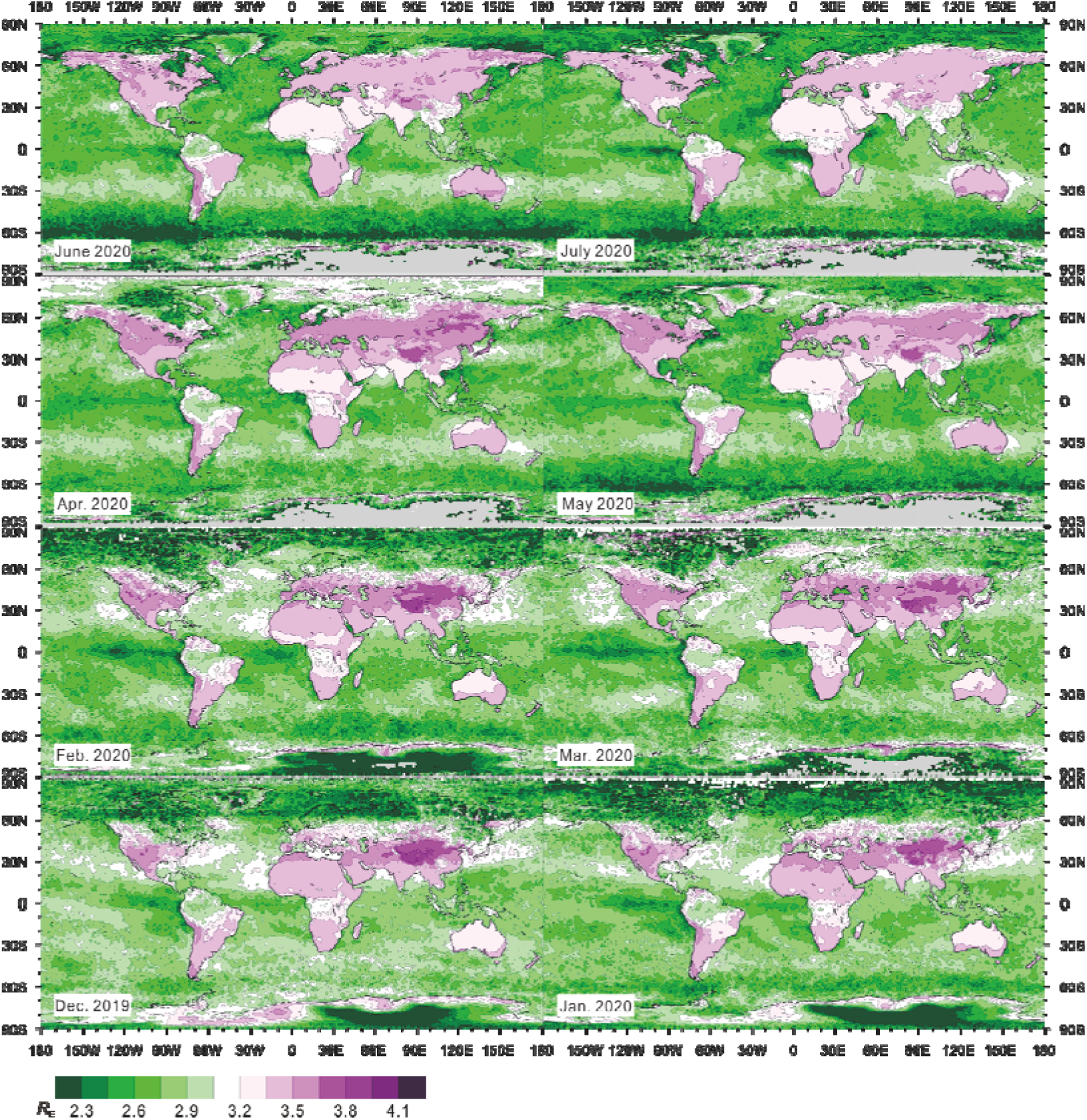
Modeled global monthly *R*_E_ value from Dec. 2019 to July 2020. The estimation from April to July was based on the mean daytime *T* and *RH* of corresponding month in 2019.

In the above *R*_E_ estimation. except for biological factors (*t*_1/2_ and *d*_i_). we only used meteorological factors. Although there are large meteorological differences between regions and dates. even within one day (causing *R*_E_ differences in a day; Figure 8). today’s meteorological satellites allow easy access to high-resolution weather data. For spread of a specific disease. therefore. *R*_E_ is a useful index for global epidemic prediction.

**Figure 8.**
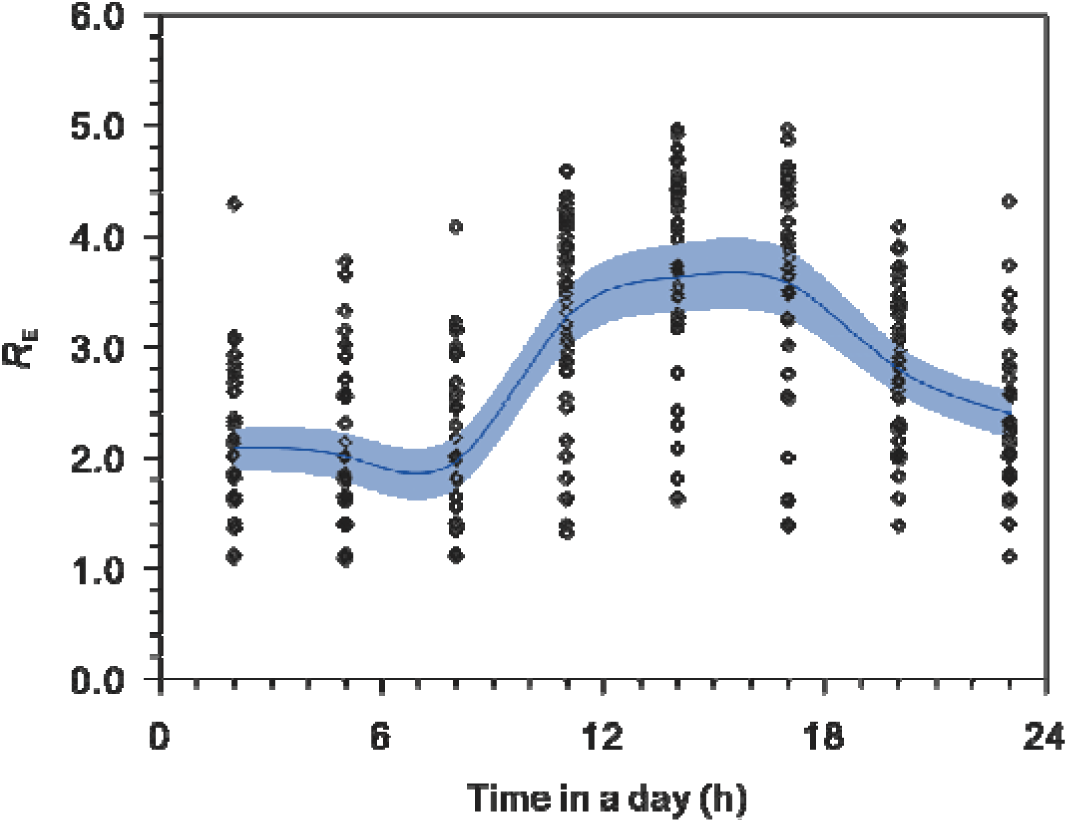
The 3-hourly modeled RE values each day in WH during the epidemic. The blue line is the average. Shaded region marks the 95% prediction envelop.

The estimation of R_E_ value is under atmospheric static and steady conditions without considering the influence of wind and airflow turbulence. Wind (>3m/s) is effective at blowing away aerosols, so it is unlikely to be infected in windy open areas (Fig. 9). Indoor airflow (i.e. caused by air conditioning and walking) can reduce the floating time of droplets and disperse viruses in the air, but also significantly increase exposure chance to viruses. Unless the MID of COVID-19 is small enough, airflow disturbance may not significantly increase the infection rate. So the modeled *R*_E_ values may be a upper limit.

**Figure 9.**
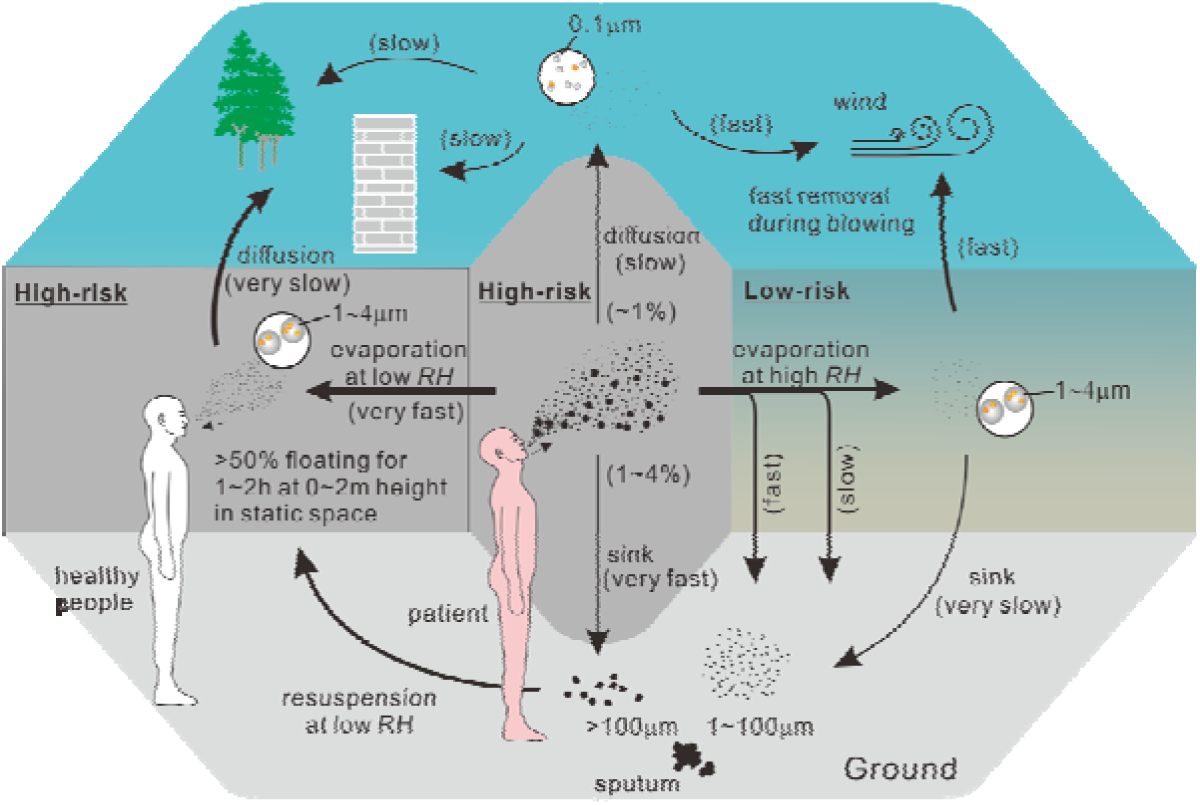
Potential ways for droplets to spread the epidemic and risk.

We noticed that the similar half lifetime of SARS-CoV-1 (caused Severe Acute Respiratory Syndrome, SARS) to SARS-CoV-2 in aerosols^20^, so the spread of the two epidemics should be consistent. But SARS disappeared in the summer of 2003, which was probably due to the infectivity only after onset of the disease, and thus more controllable than COVID-19^1^, namely, the *d*_i_ value is lower. This also confirms the importance of the control measures^12,13^.

## 4. Conclusions

In summary, we can easily obtain the global *R*_E_ value of COVID-19 of any time. The applications in WH and to global epidemic development from Feb. to Apr. are successful, making it possible to predict the global transmission of COVID-19 in the upcoming summer. Our estimation showed that warmer weather after April will help slow down the outbreak in the 30°-60°N zone, but the natural extinction of COVID-19 is less likely in summer. Northern Europe, Russia and Canada should be more alert to spread of the epidemic in May.

## Data Availability

The number of confirmed cases of coronavirus disease 2019 (COVID-19) in all countries, except China, is from Johns Hopkins CSSE (https://www.arcgis.com/apps/opsdashboard/index.html#/bda7594740fd40299423467b48e9ecf6,). The number in Chinese provinces except Hubei is from Chinese Center for Disease Control and Prevention (http://2019ncov.chinacdc.cn/2019-nCoV/,). The number in Hubei is from National Health Commission of the People’s Republic of China (http://www.nhc.gov.cn/xcs/yqtb/list_gzbd.shtml). The number of the onset in Wuhan is from Report of the WHO-China Joint Mission on Coronavirus Disease 2019 (COVID-19).
The data of monthly and 10-days average global surface temperature (T, oC) and relative humidity (RH, %) is from Geospatial Interactive Online Visualization and Analysis Infrastructure (Giovanni, https://giovanni.gsfc.nasa.gov/giovanni/, AIRS/Aqua L3 Daily Standard Physical Retrieval (AIRS-only) V006 1 oC × 1 oC). The ground station data of temperature and relative humidity is from ground observation data (http://www.weatherandclimate.info and https://www.wunderground.com/history).
Population data leaving Wuhan is from https://qianxi.baidu.com/. Population grouped by age in China, Guangzhou and Wuhan are from Chinese National Bureau of Statistics (http://www.stats.gov.cn/tjsj/ndsj/, 2017), Guangzhou Statistics Bureau (http://tjj.gz.gov.cn/pchb/dlcrkpc/, the sixth census) and Wuhan Statistics Bureau http://tjj.wuhan.gov.cn/newslist.aspx?id=2012111010461248, 2017), respectively.

## References

1. Sajadi, M.M., et al. Temperature, humidity and latitude analysis to predict potential spread and seasonality for COVID-19. SSRN http://dx.doi.org/10.2139/ssrn.3550308 (2020)

2. Yao, Y., et al. No Association of COVID-19 transmission with temperature or UV radiation in Chinese cities. Eur. Respir. J. DOI: 10.1183/13993003.00517-2020 (2020).

3. Tian, H., et al. An investigation of transmission control measures during the first 50 days of the COVID-19 epidemic in China. Science DOI:10.1126/science.abb6105 (2020).

4. Wu, J.T., Leung, L. & Leung, G.M. Nowcasting and forecasting the potential domestic and international spread of the 2019-nCoV outbreak originating in Wuhan, China: a modelling study. Lancet 395, 689–697 (2020).

5. Ahmed, F., Ahmed, N.E., Pissarides, C. & Stiglitz, J. Why inequality could spread COVID-19. Lancet Public Health. DOI: 10.1016/S2468-2667(20)30085-2 (2020).

6. Hemmes, J.H., Winkler, K.C. & Kool, S.M. Virus survival as a seasonal factor in influenza and poliomyelitis. Nature 188, 430–431(1960).

7. Heikkinen, T. & Järvinen, A. The common cold. Lancet 361, 51–59 (2003).

8. Price, R.H.M., Graham, C. & Ramalingam, S. Association between viral seasonality and meteorological factors. Sci. Rep. DOI:10.1038/s41598-018-37481-y (2019).

9. Anderson, R.M., Heesterbeek, H., Klinkenberg, D., & Hollingsworth, T.D. How will country-based mitigation measures influence the course of the COVID-19 epidemic? Lancet 395, 931–934 (2020).

10. Dao, A., et al. Signatures of aestivation and migration in Sahelian malaria mosquito populations. Nature 516, 387–390 (2014).

11. Mallapaty, S. Why does the coronavirus spread so easily between people? Nature DOI:10.1038/d41586-020-00660-x (2020).

12. Lipsitch, M., et al. Transmission dynamics and control of severe acute respiratory syndrome. Science 300, 1966–1970 (2003).

13. Riley, S., et al. Transmission dynamics of the etiological agent of SARS in Hong Kong: impact of public health interventions. Science 300, 1961–1966 (2003).

14. Shaman, J. & Kohn, M. Absolute humidity modulates influenza survival, transmission, and seasonality. Proc. Natl Acad. Sci. USA, 106, 3243–3248 (2009)

15. Ridenhour B, Kowalik JM, Shay DK. Unraveling R_0_: considerations for public health applications. Am. J. Public Health. 104, e32–41 (2014).

16. National Health Commission of the People’s Republic of China. China–WHO COVID-19 (COVID-19) joint investigation report. Feb. 29, 2020. http://www.nhc.gov.cn/iki/pqt/newlist2.shtml (in Chinese).

17. Marr, L. C., Tang, J. W., Van Mullekom, J., & Lakdawala, S. S. Mechanistic insights into the effect of humidity on airborne influenza virus survival, transmission and incidence. J. R. Soc. Interface 16, 20180298. (2019). http://dx.doi.org/10.1098/rsif.2018.0298

18. Duguid, J.P. The size and the duration of air-carriage of respiratory droplets and droplet-nuclei. J. Hyg. 4, 471–480 (1946).

19. Posada, J.A., Redrow, J. & Celik, I. A mathematical model for predicting the viability of airborne viruses. J. Virol.l Methods 164, 88–95 (2010).

20. Bushmaker, T., et al. Aerosol and Surface Stability of SARS-CoV-2 as Compared with SARS-CoV-1. N. Engl. J. Med. DOI: 10.1056/NEJMc2004973 (2020).

21. The Novel Coronavirus Pneumonia Emergency Response Epidemiology Team, CCDCP. The epidemiological characteristics of an outvreak of 2019 novel coronavirus (COVID-19) in China. Chin. J. Epidemiol. 41(2), 145–151 (2020) (in Chinese)

22. Li, Q., et al. Early transmission dynamics in Wuhan, China, of novel coronavirus–infected pneumonia. N. Engl. J. Med. 382, 1199–1207 (2020).

23. Guan, W., et al. Clinical Characteristics of Coronavirus Disease 2019 in China. N. Engl. J. Med. DOI: 10.1056/NEJMoa2002032 (2020).

24. Giardina, M. & Buffa, P. A new approach for modeling dry deposition velocity of particles. Atmos. Environ. 180, 11–22 (2018).

